# Drug treatments affecting ACE2 in COVID-19 infection: a systematic review protocol

**DOI:** 10.1101/2020.05.25.20100438

**Authors:** Hajira Dambha-Miller, Ali Albasri, Sam Hodgson, Chris Wilcox, Nazrul Islam, Shareen Khan, Paul Little, Simon J Griffin

**Affiliations:** NIHR Academic Clinical Lecturer and GP, Division of Primary Care and Population Health, University of Southampton; Clinical Pharmacist and Research Fellow, Nuffield Department of Primary Care Health Sciences, University of Oxford; NIHR Academic Clinical Fellow, Division of Primary Care and Population Health, University of Southampton; Clinical Trial Service Unit and Epidemiological Studies Unit, Nuffield Department of Population Health, University of Oxford, Oxford, United Kingdom; Medical Research Council Epidemiology Unit, University of Cambridge, Cambridge, United Kingdom; Specialist Pharmacist, Oxford University Hospitals NHS Foundation Trust; Professor of Primary Care, Division of Primary Care and Population Health, University of Southampton; Professor of General Practice, Department of Public Health and Primary Care, School of Clinical Medicine, University of Cambridge

**Keywords:** COVID-19, SARS-CoV-2, ACE2, Systematic Review

## Abstract

**Background:** The SARS-CoV-2 virus causing COVID-19 binds human angiotensin-converting enzyme 2 (ACE2) receptors in human tissues. ACE2 expression may be associated with COVID-19 infection and mortality rates. Routinely prescribed drugs which up- or down-regulate ACE2 expression are therefore of critical research interest as agents which might promote or reduce risk of COVID-19 infection in a susceptible population.

**Aim:** To review evidence on routinely prescribed drug treatments in the UK that could up- or down-regulate ACE2 and potentially affect COVID-19 infection.

**Design and setting:** Systematic review of studies published in MEDLINE, EMBASE, CINAHL, the Cochrane Library and Web of Science from inception to April 1^st^ 2020.

**Method:** A systematic review will be conducted in line with PRISMA guidelines. Inclusion criteria will be: i) assess effect of drug exposure on ACE2 level; ii) drug is included in British National Formulary (BNF) and therefore available to prescribe in UK; iii) a control, placebo or sham group is included as comparator. Exclusion criteria will be: i) ACE2 measurement in utero; ii) ACE2 measurement in children under 18 years; iii) drug not in BNF; iv) review article. Quality will be assessed using the Cochrane risk of bias tool for human studies, and the SYRCLE risk of bias tool for animal studies.

**Results:** Data will be reported in summary tables and narrative synthesis.

**Conclusion:** This systematic review will identify drug therapies which may increase or decrease ACE2 expression. This might identify medications increasing risk of COVID-19 transmission, or as targets for intervention in mitigating transmission.

## How this fits in

Clinicians, researchers and patients are increasingly interested in whether existing drug treatments may influence outcomes in COVID-19. As the binding site for SARS-CoV-2, ACE2 is of particular interest. In this systematic review, we will identify what evidence exists on effects of drugs prescribed in the UK on ACE2 levels. The findings will highlight drugs which might promote or prevent transmission of COVID-19.

## Introduction

The rapid spread of the novel, pathogenic SARS-CoV-2 (Severe Acute Respiratory Syndrome Coronavirus 2) - between late 2019 and April 2020 has escalated to a global health emergency^1^. As of April 2020 over 2 million individuals have been infected worldwide; over 150,000 people have died of the associated disease COVID-19 (COronaVirus Disease 2019)^2^. Focus on strategies to reduce viral transmission and disease mortality has intensified, including both pharmacological^3^ and non-pharmacological approaches^4,5^, alongside high speed vaccine development programmes^6^.

SARS-CoV-2 has been shown to enter cells via binding to the angiotensin-converting enzyme 2 (ACE2)^7^. The ACE2 receptor performs a broad range of roles in a variety of tissues including lung, kidney, brain and heart^8^. As part of the renin-angiotensin-aldosterone system (RAAS), ACE2 is implicated in roles including blood pressure homeostasis^9^ and downregulation of inflammation^10^. It has been hypothesises that SARS-CoV-2 may mediate effects contributing to viral sepsis and mortality through interaction with ACE2 in a variety of tissues^11^. Furthermore, differential expression of ACE2 in disease states such as hypertension may be associated with differing rates of COVID-19 infection or mortality^12^.

A key research question is whether any routinely prescribed drugs may be associated with altered ACE2 expression, and whether this altered expression may be of clinical relevance in COVID-19. For example, debate is ongoing about whether patients taking ACE inhibitors should be advised to switch drug class due to their association with increased ACE2 expression^13,14^. Conversely, medications which downregulate ACE2 transmission may be interest as part of a strategy to reduce viral transmission^15^.

In this systematic review we seek to identify what evidence exists on existing drug therapies that may increase or decrease ACE2 expression. Identifying drugs with these effects will help direct future COVID-19 research by highlighting drugs which may either prevent or promote transmission of the disease.

## Methods and Analysis

### Protocol Development

The systematic review will be conducted in line with guidance set out in the Preferred Reporting Items for Systematic Review and Meta-Analysis PRISMA statement^16^.

### Search Strategy

Systematic searches will be conducted from inception until 1/4/2020 in the following databases: MEDLINE, EMBASE, CINAHL, the Cochrane Library, and Web of Science. The detailed search strategy is provided in Table1. References of relevant reviews and articles meeting selection criteria will additionally be screened; advice will be sought from topic experts; and OpenGrey will be searched to identify additional texts. We will set no language limits or study design filters on our search.

**Table 1:** Search strategies to be used in systematic review, described by database to be searched. Terms Database.

| Terms | Database |
| --- | --- |
| (ace2[Title/Abstract]) OR ace 2[Title/Abstract]<br>Filters: English | Medline (Pubmed) |
| (ace2 or ace 2).ab. and English.lg | Embase |
| ace2 in Title Abstract Keyword OR ace-2 in Title Abstract Keyword - (Word variations have been searched) | Cochrane |
| (TS=(ace2 OR ace-2)) AND LANGUAGE: (English)<br>AND DOCUMENT TYPES: (Article) | Web of Science |

### Study selection criteria

The study inclusion criteria will be:

1. Studies measuring ACE2 levels (of either activity or gene expression)
2. Studies including a drug that is available on a UK prescription according to the British National Formulary (BNF)^17^
3. Measuring the effect of the drug against a control, placebo or sham group

Study exclusion criteria will be:

1. ACE2 measurements in utero
2. Studies in children under 18 years of age
3. The medication being studied is not available to prescribe in primary care in the UK, as assessed by inclusion in the (BNF)
4. Review articles

All methodologies meeting the above criteria, including randomised and non-randomised controlled trials and other experimental studies, will be included. Conference abstracts will be included if sufficient data can be elicited from them. Review articles will be excluded from synthesis, but their references will be screened and all appropriate studies included.

### Screening & Data Extraction

Study titles and abstracts will be reviewed for eligibility by four members of the review team (AA, HDM, CW, SH). Full text review, data extraction and quality assessment will be performed by five members of the team (AA, HDM, CW, SH, SK). Initial screening will be performed using the systematic review web app Rayyan QCRI^18^.

The following data will be extracted and verified by one of the five members of the review team (AA, HDM, CW, SH, SK): i) drug class; ii) drug name; iii) duration of treatment; iv) effect on ACE2 level (defined as upregulation or downregulation or no effect); v) model (e.g. human, mouse); vi) site of ACE2 receptor (e.g. lung, brain); vii) study design; viii) study population; ix) sample size; x) country. Given the urgency of this research question during the COVID-19 pandemic, we will extract all available information from the text but will not separately contact authors.

### Quality Assessment

Methodological quality of included animal studies will be assessed using the SYRCLE risk of bias tool (SYstematic Review Center for Laboratory animal Experimentation)^19^. This tool has been adapted from the Cochrane risk of bias tool for randomised studies^20^ to include criteria to specifically assess the quality of animal studies. The quality of human studies will be analysed separately using the Cochrane risk of bias tool.

### Summary Tables and Narrative Synthesis

Numbers of studies identified in searches and subsequently included in analysis will be demonstrated using the PRISMA flowchart in line with PRISMA guidance^16^. Extracted data will be presented in summary tables to meet the primary objective of this review.

Given the expected heterogeneity in study designs, models included in analysis (including in vivo and in vitro human and animal studies), and methods for measuring ACE2, meta-analysis via forest plots will not be appropriate. A narrative synthesis of results will be produced. Where inconsistencies are identified in the effect of a drug between studies, we will consider additional data such as materials, quality of study, and outcome measurement for potential explanatory factors.

### PPI

Patients and the public were not directly involved in the design of this study due to funding limitations. We have invited patients to help us to develop our dissemination strategy.

### Amendments

Any amendments to this protocol will be documented and communicated in the final prepared manuscript.

### Dissemination

The results of this review will be published in an open access journal to ensure free and immediate access for researchers and clinicians. Findings will also be disseminated in various media including through presentation at medical conferences and through digital platforms, including research group websites and social media.

## Discussion

### Summary

This review will deliver timely and key answers to an important question amidst the COVID-19 pandemic: do any routinely prescribed drugs up-or down-regulate ACE2 expression and therefore play a potential role in disease transmission?

### Strengths and limitations

The strengths of this review protocol include its broad search strategy; inclusion of both human and animal studies; inclusion of studies from all languages; and the intention to rapidly assess and synthesise the evidence to meet the pressing research needs of the COVID-19 pandemic. Weaknesses include anticipated heterogeneity across animal and human models which may complicate result interpretation. Furthermore, we do not intend to contact authors of papers to obtain missing data due to the need to urgently report findings amidst the current pandemic.

### Comparison to existing literature

We believe this to be the first systematic review assessing associations between drug exposure and ACE2 expression for drugs routinely prescribed in the UK.

### Implications for practice

Identification of the best evidence on these drugs might, for example, inform clinicians considering whether patients should be advised to stop ACE inhibitors^21^. Conversely, drugs which are associated with ACE2 downregulation might represent a therapeutic strategy to reduce viral transmission and disease spread.

## Data Availability

All data are included within the manuscript ie search terms

## Acknowledgements

None

## Ethical Approval

Not required

## Data sharing

No other data available

## Author Contribution

HDM contributed to the design of the study, wrote the analysis plan, conducted the analysis, and revised the paper. SH contributed to the design of the study, drafted and revised the paper. PL, AA, CW, NI and SK and SG contributed to the design of the study and revised the paper. HDM is guarantor. All authors will be involved in delivery of the systematic review in line with this protocol.

## Patient and Public Involvement

It was not possible to involve patients or the public in the design or conduct of our work due to the rapid timelines, but we have invited PPI representatives to help us with drafting a lay summary and in the dissemination of our findings.

## Supplementary Data

Nil

